# Early Pregnancy DNA Methylation Signatures as Predictors of Antenatal Depressive Symptoms: A longitudinal study of DNA methylation changes

**DOI:** 10.64898/2026.02.04.26345531

**Authors:** Kuppan Gokulakrishnan, Chinnasamy Thirumoorthy, Kuldeep Sharma, Mohan Deepa, Venkatesh Ulagamadesan, Bettadapura N Srikumar, Bhaskarapillai Binukumar, Uma Ram, Ranjit Mohan Anjana, Muthuswamy Balasubramanyam, Viswanathan Mohan, Ponnusamy Saravanan

## Abstract

**Background:** Antenatal depressive symptoms (ADS) are common and underdiagnosed, particularly in low- and middle-income countries, and are associated with adverse maternal and offspring outcomes. Current screening relies on subjective symptom reporting, limiting early identification and prevention. Epigenetic modifications, particularly DNA methylation, offer a promising avenue for objective, early biomarkers of depression risk during pregnancy.

**Methods:** In this nested case–control design within the STRiDE (Stratification of Risk of Diabetes in Early Pregnancy study) prospective cohort, 189 pregnant women with no depressive symptoms in early pregnancy (<16 weeks of gestation. PHQ-9 ≤4) were followed longitudinally. 89 ADS individuals were identified by the emergence of depressive symptoms at 24–28 weeks of gestation (PHQ-9 ≥5), while 100 women remained symptom-free (Controls). Genome-wide DNA methylation profiling of early-pregnancy peripheral blood was performed using the Illumina EPIC 850K array. Epigenome-wide association analyses were combined with machine-learning approaches to identify predictive CpG panels. Model robustness was assessed using bootstrap validation, and a methylation risk score (MRS) was constructed. Functional enrichment analyses were conducted to explore biological pathways.

**Results:** Epigenome-wide analysis identified 2,447 differentially methylated positions associated with subsequent ADS. A robust panel of 10 CpGs in early pregnancy predicted later ADS with excellent performance (testing AUC=0.99. bootstrap-validated AUC=0.91), independent of maternal risk factors. The MRS markedly outperformed traditional clinical predictors (AUC=0.94) and further improved prediction when combined with maternal characteristics (AUC=0.95). ADS-associated methylation changes were enriched in neurodevelopmental, synaptic, immune, and metabolic signaling pathways. Limited concordance with placental methylation suggested maternal-specific epigenetic regulation.

**Conclusions:** Early-pregnancy DNA methylation signatures can predict antenatal depressive symptoms before clinical onset. This blood-based 10-CpG biomarker panel offers a biologically informed and objective tool for early risk stratification, with the potential to enable preventive interventions and enhance perinatal mental health care, particularly in resource-constrained settings.

## Introduction

Depression during pregnancy represents a significant public health concern, with global prevalence estimates ranging from 15% to 65% (Dadi et al., 2020). Rates are notably higher in low- and middle-income countries compared to high-income settings. In India, reported prevalence varies from 9% to 36%, depending on regional and methodological differences (Ajinkya et al., 2013; Jyothi Kantipudi et al., 2020). Despite its high burden, antenatal depression often remains underrecognized and undertreated, particularly in resource-limited settings (Dadi et al., 2020). Antenatal depression is associated with a higher risk of obstetric complications such as gestational diabetes mellitus, hypertension, and preeclampsia, as well as adverse birth and child outcomes, including low birth weight, preterm delivery, developmental delays, and disrupted neurodevelopmental trajectories (Gelaye et al., 2016; Gentile, 2017; Herba et al., 2016). Furthermore, maternal depression during pregnancy elevates the risk of psychiatric disorders in offspring later in life (Pietikäinen et al., 2020).

Evidence suggests that the second and early third trimesters are particularly sensitive periods during which exposure to stressful life events can exert long-lasting biological and psychological impacts on the offspring (O’Connor et al., 1995; Rogers et al., 2020; Van den Bergh et al., 2008). Untreated antenatal depression has been shown to be a strong predictor of postpartum depression, with long-term implications for maternal–infant bonding, attachment, and early childhood development (Luciano et al., 2022; Wang et al., 2021). However, current screening methods primarily rely on self-reported symptom assessments, which can be subjective and influenced by sociocultural stigma, underreporting, or variability in clinical expression. This underscores the urgent need for objective, biologically based tools that can predict the onset of depressive symptoms during pregnancy before clinical manifestation.

Epigenetic modifications, particularly DNA methylation, may serve as the molecular link between environmental influences and gene expression, and have emerged as promising early biological markers for mood disorders during pregnancy (Campagna et al., 2021; Skinner, 2024; Thirumoorthy et al., 2025b). DNA methylation changes have been associated with depression in both clinical and population-based studies, suggesting that peripheral blood methylation profiles may reflect stress-related biological changes in the central nervous system (Løkhammer et al., 2022; Nie et al., 2022). Recent studies have demonstrated that DNA methylation alterations in maternal blood and placental tissue were associated with depressive symptoms during the second and third trimesters of pregnancy (Kee et al., 2022; Lund et al., 2021; Tesfaye et al., n.d.; Viuff et al., 2018). These findings suggest that DNA methylation signatures may serve as early molecular indicators of vulnerability to mood disturbances. Most existing studies are cross-sectional and focus on methylation profiles after the onset of depressive symptoms, offering limited insight into early molecular changes that precede clinical presentation. Moreover, no laboratory-based diagnostic test for depression currently exists, advances in machine learning and deep learning studies indicate that DNA methylation profiles can be leveraged as predictive biomarkers for neuropsychiatric conditions, including depression (Sokolov and Schiöth, 2024).

Despite growing evidence, studies exploring epigenetic biomarkers of antenatal depression remain limited, especially in low- and middle-income countries. India, with its unique sociocultural stressors, nutritional disparities, and genetic diversity, provides a distinct context in which the biological correlates of depression may differ from those observed in Western populations. In this context, our recent studies indicate that early trimester telomere length and mitochondrial DNA copy number could be potential predictive biomarkers for predicting depressive symptoms(Thirumoorthy et al., 2025a). Additionally, we have shown that DNA methylation signatures in maternal blood leukocytes at the time of depression diagnosis can predict subsequent postpartum depression (Thirumoorthy et al., 2025b). These findings support the notion that molecular alterations during pregnancy may reflect the biological impact of psychological stress and serve as early indicators of adverse mental health outcomes. Building upon the above insights, the present longitudinal study aims to fill the gaps by investigating whether DNA methylation signatures in early pregnancy, alone or together with clinical factors, can predict the development of antenatal depressive symptoms in later trimesters, thereby offering a valuable new tool for antenatal depression risk stratification and prevention

## Materials and Methods

### Study Design and Participants

This nested case–control study included 189 women enrolled in the prospective STratification of Risk of Diabetes in Early Pregnancy (STRiDE) study (Saravanan et al., 2024). The study was approved by the Institutional Ethics Committees of the National Institute of Mental Health and Neuro Sciences (NIMHANS), Bengaluru, and the Madras Diabetes Research Foundation (MDRF), Chennai, India. Written informed consent was obtained from all participants prior to enrolment.

All 189 pregnant women had a Patient Health Questionnaire-9 (PHQ-9) score of ≤4 during early pregnancy (<16 weeks of gestation). Participants with a prior or current psychiatric illness at recruitment were excluded. Assessment of psychiatric history was conducted through a comprehensive clinical interview by trained mental health professionals, supplemented by review of available medical records. Women with any documented history of psychiatric disorders including major depression, bipolar disorder, psychosis or those on psychotropic medications (antidepressants, antipsychotics, or mood stabilizers) were excluded to minimize confounding from prior psychiatric treatment or medication effects on DNA methylation.

### Classification of study participants and DNA methylation profiling

Participants were stratified into two groups - antenatal depressive symptoms (ADS) (n = 89) and controls (n = 100) based on PHQ-9 scores at the oral glucose tolerance test (OGTT) visit (24–28 weeks of gestation). The PHQ-9 assessment was performed at two points: early pregnancy and at the OGTT visit. Participants with PHQ-9 ≤4 at both visits were classified as controls, whereas those with PHQ-9 ≤4 during early pregnancy but ≥5 at the OGTT visit were classified as ADS.

DNA methylation profiling was performed on 189 blood samples collected at each visit (early pregnancy and OGTT), as well as on a subset of 107 placental samples.

### Anthropometric and Clinical Measurements

Trained personnel obtained anthropometric and clinical data using standardized procedures. Weight, height, and waist circumference were measured, and body mass index (BMI) was calculated as weight (kg) divided by height (m^2^). Blood pressure was measured in the right arm while seated, using an OMRON electronic sphygmomanometer (Omron Corporation, Tokyo, Japan). Two readings were taken 5 minutes apart, and the average value was recorded.

### Evaluation of Antenatal Depressive Symptoms

Depressive symptoms were evaluated during early pregnancy and again at the OGTT visit using the PHQ-9 questionnaire. The PHQ-9 is a validated screening tool for depressive symptoms based on the Diagnostic and Statistical Manual of Mental Disorders, Fourth Edition (DSM-IV) criteria for major depressive disorders. It has demonstrated robust psychometric properties and high sensitivity and specificity across diverse populations, including pregnant and postpartum women (Hirschtritt and Kroenke, 2017; Rathod et al., 2018; van Heyningen et al., 2018; Wang et al., 2021). Previous validation studies in Indian populations identified a PHQ-9 cutoff score of ≥5 as optimal for identifying depressive symptoms (Ali et al., 2020; Prabu et al., 2020; Rathod et al., 2018). Therefore, a PHQ-9 score ≥5 was used in this study to define the presence of depressive symptoms.

### Sample Collection and DNA Extraction

#### Peripheral Blood Samples

Fasting peripheral blood samples were collected from all participants (n = 189) at two time points - early pregnancy and at the OGTT visit - following an overnight fast. Genomic DNA was extracted from peripheral blood using the phenol–chloroform method. DNA concentration and purity were assessed using a NanoDrop OneC spectrophotometer (Thermo Fisher Scientific, USA), and DNA integrity was verified by electrophoresis on a 0.8% agarose gel.

#### Placental Tissue Samples

Following delivery, placental tissue samples were obtained from a subset of the same participants (n = 107), including 53 ADS and 54 controls. Term placentas were sampled from three sites within the inner quadrant of the fetal side, avoiding areas of infarction, hematoma, or visible damage. Tissue sections (∼1 × 1 inch) were rinsed thoroughly with phosphate-buffered saline (PBS) to remove residual blood, flash-frozen in liquid nitrogen, and stored at −80°C until further processing.

Placental genomic DNA was isolated using the QIAamp DNA Mini Kit (Qiagen, Germany) according to the manufacturer’s instructions. DNA purity and concentration were measured using a NanoDrop OneC spectrophotometer, and integrity was confirmed by visualization on a 0.8% agarose gel.

#### Bisulfite conversion, DNA methylation profiling & data analysis

Bisulfite conversion was performed using the EZ DNA Methylation Kit from Zymo Research (USA) and subsequently analyzed with the HumanMethylationEPIC 850k BeadChip assays (Illumina, San Diego, CA, USA), following the manufacturer’s instructions and the beadchips were scanned on an iScan reader (Illumina). To minimize potential batch effects, both case and control samples were processed and ran concurrently on the same BeadChip.

CpGs with poor signal quality (detection p-value >0.05), those located on the X and Y chromosomes, common SNPs, and cross-reactive probes were excluded from the analysis. This resulted in a final dataset comprising 189 samples and 537,196 CpGs for EWAS. Prior to conducting EWAS, data normalization was performed using the quantile normalization method. EWAS was carried out, using the lmFit() function from the limma package. This function calculates the p-value for differential methylation using a linear model, with the CpGs (beta values) as the dependent variable and the ADS (Present/Absent) as the independent variable.

The effect of ADS was adjusted for the cell type proportions as well. Cell type proportions were extracted for six cell types: Granulocytes, CD4T, CD8T, B cell, Monocytes, and natural killer (NK) cells, using the EpiDISH package in R (Houseman et al., 2014, 2012; Zheng et al., 2020). As anticipated for blood samples, we found that the cell type proportions were correlated, with coefficients ranging from −0.53 to 0.25. Particularly between Granulocytes and CD8T, the correlation coefficient was high, −0.53. Due to the observed moderate to strong correlations among the cell type proportions, we computed variance inflation factors (VIFs) to assess the potential violation of the assumption of no multicollinearity. As expected, the VIFs for CD8T, B cells, and Granulocytes were notably high. Therefore, we employed a stepwise removal process, and excluded CD8T, and the VIF values approached to 1. Consequently, only CD4+ T cells, NK cells, B cells, Monocytes, and granulocytes were retained for adjustment in the EWAS. The output primarily contains fold change, p-value, and adjusted p-value (Benjamini-Hochberg).

The output included key parameters such as fold change, p-value, and Benjamin-Hochberg adjusted p-value as FDR<0.01 to identify differentially methylated CpGs, resulting in a total of 359 CpGs. Ultimately, we selected 275 CpG sites that were annotated to genes for further data analysis. We used Boruta SHAP for feature selection and selected 60 CpGs as an important feature from 275 CpGs. Radial Kernel Support Vector Machines (SVM) - were applied to construct biomarker panels using the CpG lists derived from the above feature selection methods as input variables. The panel construction employed a forward stepwise strategy aimed at optimizing the AUC, incrementally adding significant CpGs one by one to enhance model performance. The overall framework for developing these biomarker panels is illustrated in **Supplementary Table 1**.

Ten iterations were conducted by randomizing the samples in the training (67%) and testing (33%) datasets. Panels of varying sizes were constructed, starting with a single CpG and expanding to ten CpGs per panel, culminating in ten unique sets of panels for each scenario, all optimized for AUC in each iteration. The stability of each biomarker panel was evaluated through bootstrap validation, comprising 1000 runs. This validation process generated aggregated metrics, including AUC, sensitivity, specificity, and accuracy, along with their corresponding standard deviations (SD) from the 1000 iterations. The panel exhibiting the lowest SD was identified as the most robust model for each scenario, as a lower SD indicates enhanced model stability. This systematic approach resulted in a compilation of the best biomarker panel for each scenario. The final panel was selected based on the highest AUC among the six panels identified.

All analyses were executed using R (version 4.3.2), employing the “train” function from the caret package to implement SVM classifier. For the SVM model parameters, C (the margin between the classifier line and support vectors) and sigma (degree of non-linearity), were automatically tuned through 10-fold cross-validation with five repeats.

#### Prediction of GDM and ADS using methylation risk scores (MRS)

Our weighted Methylation Risk Score (MRS) was developed using a panel of CpG sites identified by machine learning classifiers for the prediction of ADS. The weighted logistic regression was used to estimate the predicted probabilities according to the CpG panels, where the MRS was estimated to be the total of the regression coefficients of the chosen 10 CpGs. The entire dataset was fitted with weighted logistic regression models to determine the ADS as the outcome, the MRS as the primary predictor, and maternal risk factors as covariates. The model was: MRS and maternal risk factors. The individual predicted risks were calculated, and the performance of the models was analysed through the ROC curve.

#### Functional enrichment GO and KEGG analysis

Gene Ontology (GO) enrichment analysis and Kyoto Encyclopedia of Genes and Genomes (KEGG) pathway enrichment analysis for the 1906 CpGs (FDR<0.05) and their annotated genes were performed using the SRplot online tool. For both the GO and KEGG analyses, the top 10 pathways were selected for further examination. A significant threshold of a false discovery rate (FDR) of less than 0.05 was established to identify biologically relevant pathways and processes associated with the DMRs.

#### Statistical Analysis

Numeric variables were summarized with mean ± SD for continuous variables and n (%) for categorical variables. The Kolmogorov–Smirnov test confirmed normality, allowing for parametric tests due to the normally distributed nature of the data. Continuous variable averages were compared using the independent t-test, while proportions were assessed using the Chi-square test or Fisher’s exact test as appropriate. Spearman correlation analysis was performed to explore the association between the best panel of CpGs at early pregnancy and PHQ-9 scores at OGTT visit. Logistic regression model’s predictive performance was assessed by plotting the receiver operating characteristic curve (ROC), from which AUC with 95% CI was calculated as the primary predictive measure. Statistical analyses were conducted using R (Version 4.3.2), with p-values <0.05 deemed statistically significant.

## Results

### Clinical and biochemical characteristics of the study participants

The demographic and clinical data of the study participants during early pregnancy, the OGTT visit, and delivery were shown in **Table 1**. The mean gestational age at early pregnancy was 10 weeks. Among the 189 study participants, 47% had ADS, and the proportion of GDM was similar across the study groups. The important baseline characteristics in early pregnancy, such as PHQ-9 score, maternal age, pre-pregnancy BMI, gestational age at recruitment, body fat percentage and blood pressure, were not significantly different between groups. The ADS individuals had significantly higher PHQ-9 score than the controls at the OGTT visit (p < 0.01). There were no significant differences between the ADS and control groups in the delivery outcomes and mode of delivery, except for birth weight.

**Table 1:** Clinical and biochemical characteristics at different visits of pregnancy.

| Variables | Controls<br>(Normal at both early pregnancy<br>& OGTT visit) (n=100) | ADS<br>(Normal at early pregnancy & become ADS<br>at OGTT visit) (n=89) |
| --- | --- | --- |
| **Early pregnancy (<16 weeks)** |  |  |
| Age (years) ^ | 27.4 ± 3.6 | 26.9 ± 3.1 |
| Pre-pregnancy body mass index (kg/m<sup>2</sup>) ^ | 24.0 ± 3.8 | 24.5 ± 4.1 |
| Gestational Age (weeks) inclusion ^ | 10.2 ± 3.0 | 9.9 ± 2.8 |
| Parity |  |  |
| First pregnancy | 74 (74%) | 75 (84%) |
| Second pregnancy | 23 (23%) | 14 (16%) |
| Third pregnancy | 3 (3%) | 0 |
| Body fat (%) ^ | 35.2 ± 6.0 | 36.2 ± 5.8 |
| Systolic Blood Pressure (mm Hg) ^ | 103.9 ± 10.4 | 102.2 ± 9.3 |
| Diastolic Blood Pressure (mm Hg) ^ | 68.7 ± 7.9 | 67.8 ± 7.1 |
| Family history of type 2 diabetes n [%] | 38 [20.1%] | 39 [20.6%] |
| PHQ-9 Depression Score ^ | 2.8 ± 1.4 | 3 ± 1.4 |
| Socio Economic Status (SES) n [%] |  |  |
| Lower class | 6 (6%) | 7 (7.9%) |
| Middle class | 63 (63%) | 55 (61.8%) |
| Upper class | 31 (31%) | 27 (30.3%) |
| **OGTT visit (24-28 weeks)** |  |  |
| Body mass index (kg/m<sup>2</sup>) ^ | 26.0 ± 3.7 | 26.7 ± 4.1 |
| Body fat (%) ^ | 39 ± 7.1 | 37.2 ± 6.3 |
| Systolic Blood Pressure (mm Hg) ^ | 104 ± 11 | 103 ± 11 |
| Diastolic Blood Pressure (mm Hg) ^ | 68 ± 8 | 67 ± 6 |
| PHQ-9 Depression Score ^ | 1.8 ± 1.6 | 8.3 ± 2.8** |
| Gestational Diabetes Mellitus n [%] | 50 [50%] | 39 [44%] |
| **Delivery Details** |  |  |
| Gestational age at delivery [weeks] ^ | 38.9 ± 1.9 | 39.3 ± 1.4 |
| Spontaneous n [%] | 48 [48%] | 45 [50.6%] |
| Instrumental n [%] | 13 [13%] | 5 [5.6%] |
| LSCS n [%] | 39 [39%] | 37 [41.6%] |
| Birthweight (grams)<sup>^</sup> | 2890.7 ± 443.5 | 3022.1 ± 411.3* |
| Child Gender-Male n [%] | 53 [53%] | 43 [48%] |
| Macrosomia n [%] | 7 [7%] | 13 [15%] |
| LGA n [%] | 1 [1%] | 3 [3.3%] |
| SGA n [%] | 20 [20%] | 16 [18%] |

<sup>^</sup>Data presented as mean ± SD; \*p < 0.05, \*\*p < 0.001, compared to controls; PHQ – Patient Health Questionnaire; OGTT – Oral Glucose Tolerance Test;; LSCS - Lower Segment Caesarean Section; LGA-large for gestational age; SGA- small for gestational age.

### Developing an early DNA Methylation-based biomarker panel for ADS prediction

We first evaluated whether DNA methylation patterns during early pregnancy could be used to predict ADS. An epigenome-wide association study (EWAS) was conducted, and after quality control and data preprocessing, 5,37,187 DNA methylation probes from 89 ADS and 100 controls were retained for further analysis. The EWAS analysis identified 2447 differentially methylated positions (DMPs) with FDR < 0.05, including 359 CpGs with FDR < 0.01. Hyper and hypomethylated CpGs associated with ADS at different thresholds were shown in the Volcano plot **(Figure 1a)**. We selected 275 CpGs which were annotated to the known genes for further analysis.

**Figure 1:**
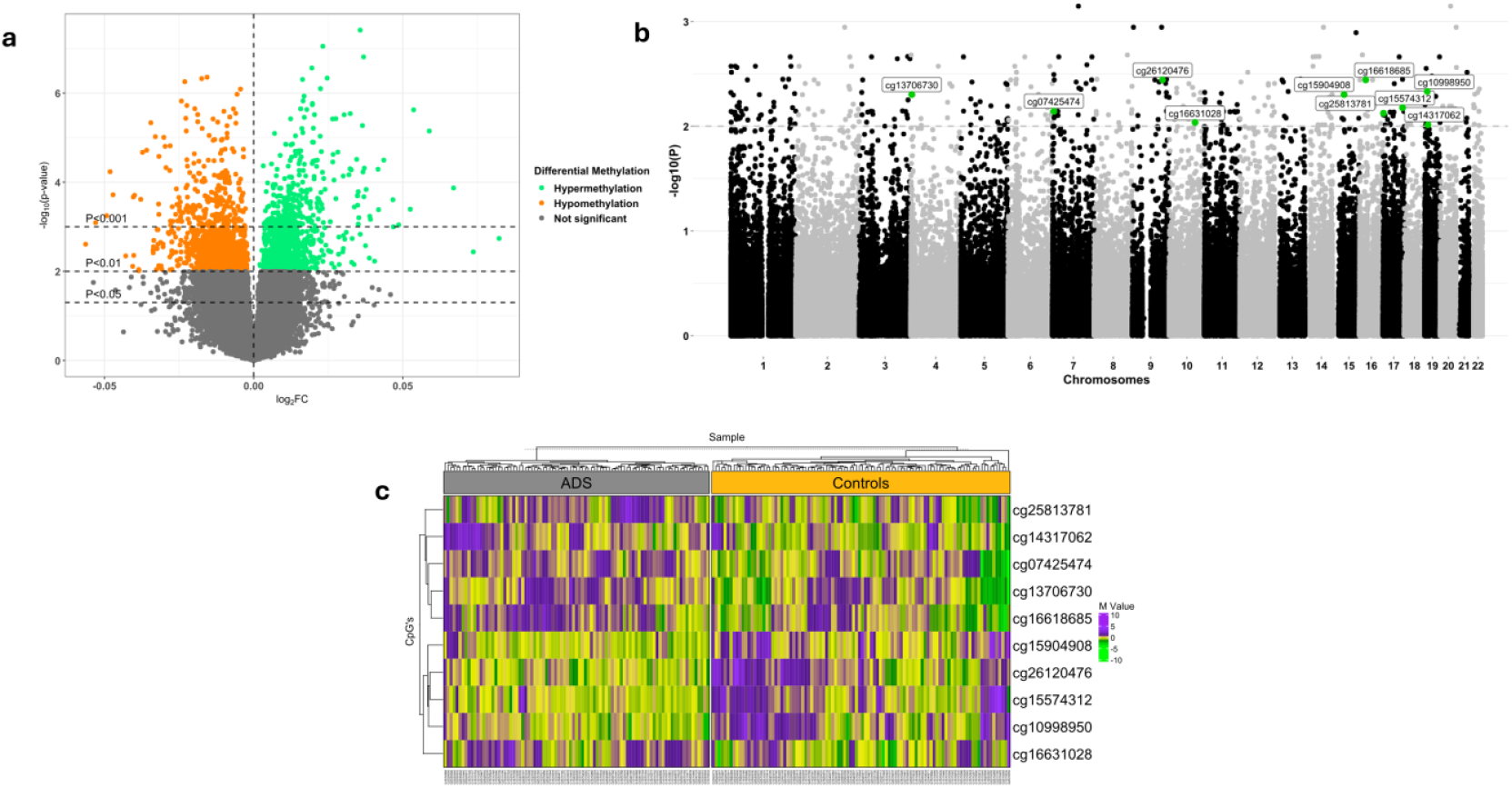
Epigenome-wide DNA methylation differences between ADS and controls in early pregnancy. (a) Volcano plot showing differentially methylated CpG sites (DMPs) between antenatal depression (ADS) and controls. Gray dots represent non-significant CpGs, while green and orange dots indicate significantly hypermethylated and hypomethylated CpGs in ADS, respectively (threshold: −log10(FDR) > 2). The x-axis represents log2 fold change, and the y-axis represents −log10(FDR). (b) Manhattan plot illustrating the chromosomal distribution of CpG sites with significant differential methylation between ADS and controls. The x-axis denotes chromosomal positions, and the y-axis represents −log10(p-values). The dotted horizontal line indicates the genome-wide significance threshold (FDR-adjusted p < 0.01). Ten CpGs identified as robust biomarkers by the SVM model are highlighted in green and labeled. (c) Heatmap depicting methylation patterns of the 10 selected DMPs in controls and ADS, demonstrating clear group-wise separation.

A radial kernel SVM machine learning model was employed to construct a predictive model for identifying a panel of CpG biomarkers that could predict ADS during early pregnancy. The SVM model yielded a panel of 10 CpGs that could predict the ADS in early pregnancy with an AUC of 99%, sensitivity of 97% and specificity of 99% in the testing data set. Bootstrap validation confirmed the robustness of this panel, with an AUC of 91%, a sensitivity of 83%, a specificity of 82%, and an accuracy of 82%. **Table 2** shows the detailed performance measures of the panel of 10 CpGs in the training, testing, and validation datasets. Combined, these results indicate that EWAS and machine learning classifier found an effective 10-CpG panel with excellent predictive accuracy for ADS in early pregnancy.

**Table 2:**
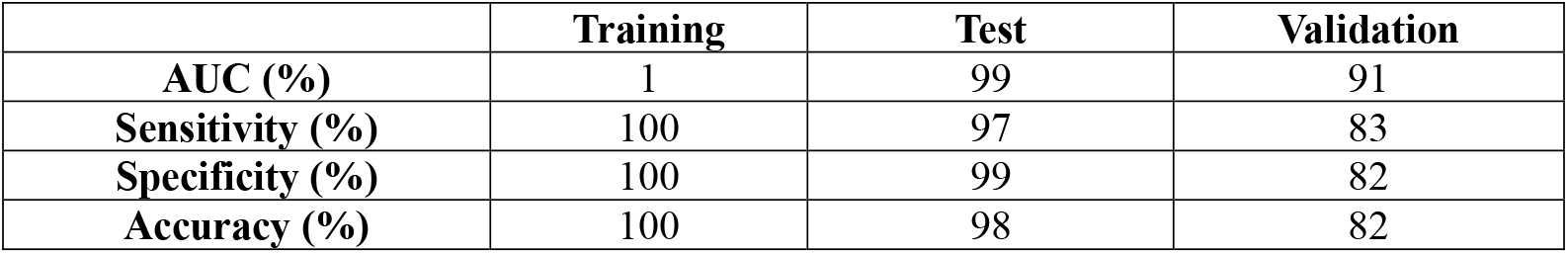
Performance metrics of the 10 CpGs biomarker panel in the Training, Test and Validation datasets.

The panel of 10 CpGs, their annotated genes and the chromosome-wide distribution were shown in the Manhattan plot **(Figure 1b)**. The heatmap shows the 10 CpGs methylation patterns between the ADS and controls **(Figure 1c)**. The 10 CpGs, their annotated genes, chromosomal location and CpG IDs were given in **Table 3**. Among 10 CpGs, six CpGs showed significantly higher methylation levels (p<0.0001) and rest four showed lower methylation levels (p<0.0001) in ADS compared to controls **(Figure 2a)**. Moreover, the feature importance analysis of 10 CpGs utilizing SHAP values indicated that the most significant CpGs for ADS prediction were cg07425474 and cg14317062 **(Figure 2b and 2c)**. Interestingly, the CpG (cg07425474) and its annotated gene (KIAA0415) were shown to be involved in the disruption of synaptic maturation and causes mood disorder-like behaviour in the mouse model (He et al., 2025).

**Table 3:** Details of best performed biomarker panel (10 CpGs) for predicting ADS using SVM Model.

| CpGs | P.Value | FDR | CHR | MAPINFO | gene | cgi | feat.cgi |
| --- | --- | --- | --- | --- | --- | --- | --- |
| cg16618685 | 4.64E-07 | 0.003608 | 16 | 23608722 | NDUFAB1 | shore | TSS1500-shore |
| cg26120476 | 4.91E-07 | 0.003608 | 9 | 1.16E+08 | SNX30 | island | TSS200-island |
| cg10998950 | 9.64E-07 | 0.004661 | 19 | 9945815 | PIN1 | island | TSS200-island |
| cg15904908 | 1.2E-06 | 0.004993 | 15 | 43213022 | TTBK2 | island | TSS200-island |
| cg13706730 | 1.22E-06 | 0.004993 | 4 | 3326208 | RGS12 | opensea | Body-opensea |
| cg15574312 | 2.24E-06 | 0.006652 | 17 | 74533845 | CYGB | island | TSS200-island |
| cg07425474 | 2.78E-06 | 0.007208 | 7 | 4825336 | KIAA0415 | island | Body-island |
| cg25813781 | 3.33E-06 | 0.00752 | 17 | 1327749 | CRK | opensea | Body-opensea |
| cg16631028 | 5.74E-06 | 0.009232 | 10 | 99204975 | EXOSC1 | shore | Body-shore |
| cg14317062 | 6.33E-06 | 0.009698 | 19 | 12777503 | MORG1 | island | TSS200-island |

**Figure 2:**
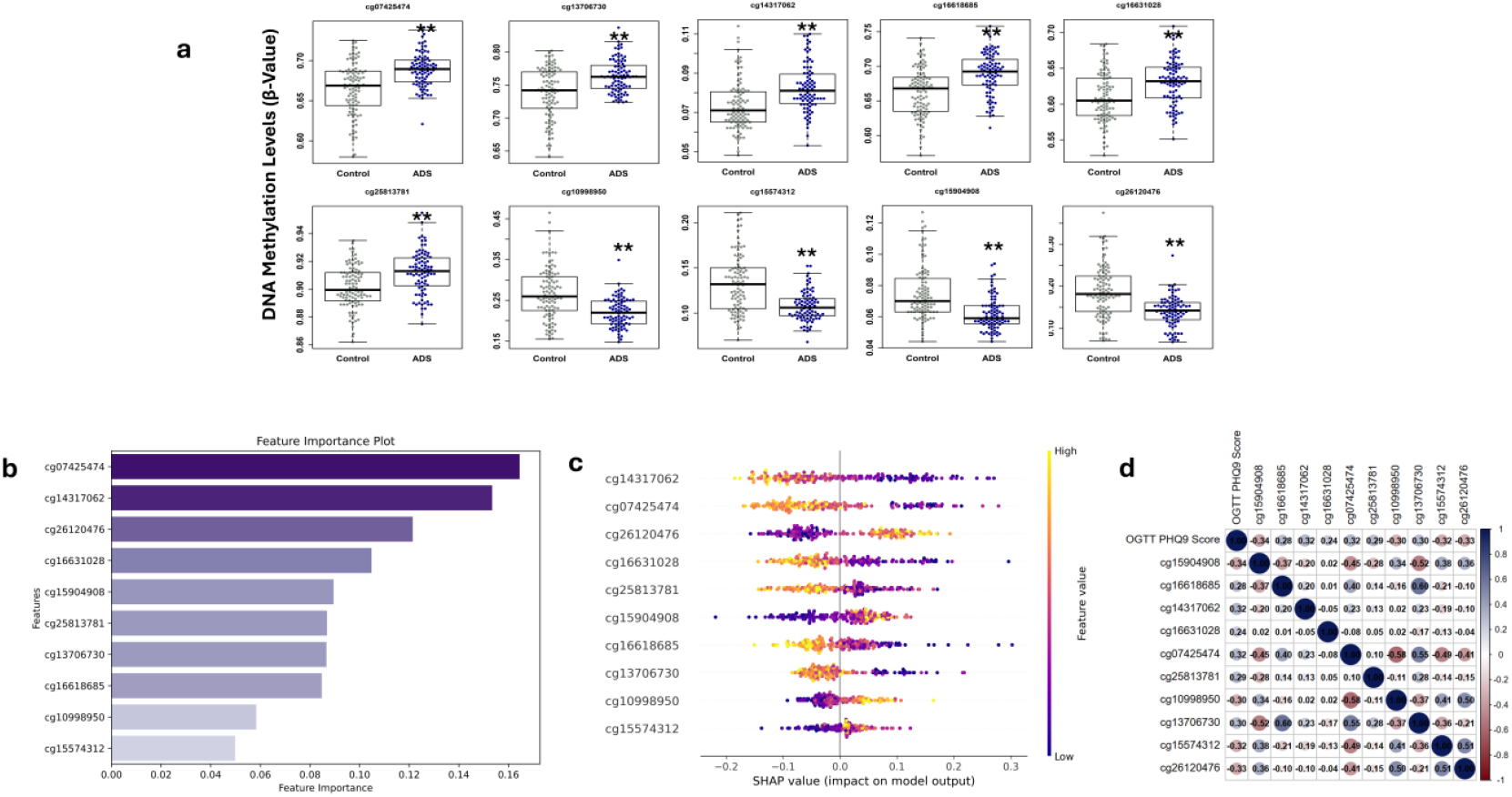
Contribution of the 10-CpG panel to ADS classification and SHAP interpretation. (a) DNA methylation levels of the top-performing 10 CpGs in controls and ADS during early pregnancy. Bar graphs represent mean ± SD. **p < 0.001 versus controls. (b) Feature importance ranking of the 10-CpG panel within the classification model. (c) SHAP summary plot showing the impact of individual CpGs on ADS classification. SHAP values on the x-axis indicate log-odds of ADS diagnosis; positive values indicate higher likelihood of ADS, while negative values indicate higher likelihood of controls. (d) Pearson correlation analysis between methylation levels of the 10 CpGs in early pregnancy and PHQ-9 scores at the OGTT visit. Color scale represents correlation strength (blue, positive; white, no correlation; red, negative), with corresponding r values shown in each cell.

The correlation analysis revealed a significant correlation (p < 0.05) between the methylation levels of the 10 CpGs during early pregnancy and PHQ-9 scores at the OGTT visit. Out of 10 CpGs, six of the CpGs demonstrated positive correlations, whereas four CpGs demonstrated negative correlations **(Figure 2d)**.

### ADS prediction by combining maternal risk factors and a panel of 10 CpGs

A relative risk (RR) analysis was conducted to determine the risk of ADS in relation to CpG methylation levels. Out of 10 identified CpGs, six showed significant positive association (p < 0.001), with higher methylation levels being associated with an increased risk of ADS: cg07425474 [RR: 1.81, 95% CI: 1.52–2.16], cg13706730 [1.61: 1.40–1.85], cg14317062 [1.40: 1.23–1.59], cg16618685 [1.58: 1.35–1.86], cg16631028 [1.36: 1.16–1.59], and cg25813781 [1.44: 1.25–1.65]. Four CpGs demonstrated an inverse association, where lower methylation levels were associated with higher risk of ADS: cg10998950 [0.61: 0.52–0.72], cg15574312 [RR: 0.55, 95% CI: 0.43–0.70], cg15904908 [0.51: 0.41–0.65] and cg26120476 [0.59: 0.50– 0.69]. Importantly, these associations remained significant after adjusting for maternal risk factors: age, pre-pregnancy BMI, gestational age, parity, child sex, and GDM status **(Figure 3a)**. Collectively, these results suggest that the panel of 10 CpGs predict ADS regardless of maternal risk factors.

**Figure 3:**
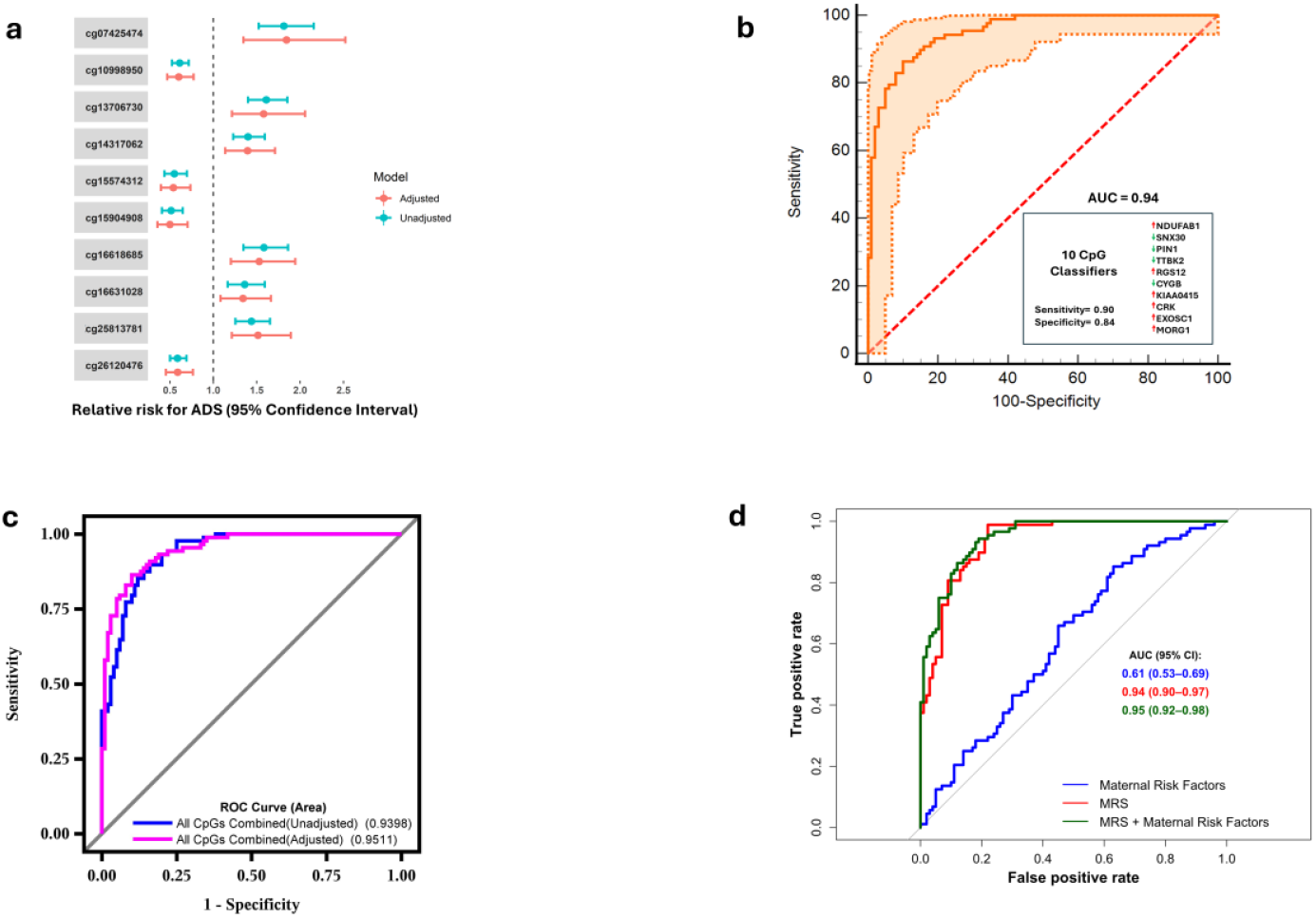
Predictive performance of the 10-CpG panel for ADS. (a) Forest plot displaying unadjusted and adjusted relative risk (RR) estimates of the 10 CpGs for ADS prediction, shown with 95% confidence intervals. (b) Receiver operating characteristic (ROC) curve assessing the discriminatory performance of the 10-CpG panel in distinguishing ADS from controls. (c) ROC analysis of the CpG panel adjusted for maternal age, pre-pregnancy BMI, PHQ-9 score, and presence of gestational diabetes mellitus (GDM). (d) ROC curves comparing the predictive performance of the methylation risk score (MRS) alone and in combination with maternal risk factors.

We conducted additional analyses of the model’s predictive performance of the 10-CpG biomarker panel for ADS prediction using ROC. The predictive performance of the panel showed an unadjusted AUC of 0.94 with sensitivity of 87%, specificity of 84%, accuracy of 86%, **(Supplementary Table 2)** and the AUC was improved to 95% after adjusting for maternal age, pre-pregnancy BMI, gestational age, parity, child sex and GDM status **(Figure 3b and 3c)**. These findings demonstrate the strong predictive utility of the 10-CpG biomarker panel, which improved after accounting for maternal risk factors.

### Development of Methylation Risk Score

The methylation risk score (MRS) was constructed as a weighted sum of the selected CpGs, with weights corresponding to the regression coefficients obtained from the binary logistic regression model. We first examined the MRS for maternal risk factors alone, which showed an AUC of 61% for ADS prediction. Initially, the MRS has demonstrated significantly greater performance, with an AUC of 94%. Following the addition of the maternal risk factors with MRS improved the AUC to 95% **(Figure 3d)**. The overall performance of these models revealed the strength of the identified CpG markers in predicting ADS during early pregnancy, regardless of the prediction methodology used.

To facilitate the use of the MRS in conjunction with maternal risk factors as an epigenetic screening test for predicting ADS, an optimal cutoff value was established to calculate predictive parameters. At the conventional probability threshold of 0.5, for MRS, the model achieved a sensitivity of 88% and a specificity of 85%, corresponding to a high Yoden index (J = 0.73). This indicated a favourable balance between sensitivity and specificity, supporting the interpretation that a predicted probability above 0.5 can be considered a reliable indicator of ADS at the individual level.

### Temporal stability and diagnostic performance of the early pregnancy 10-CpG methylation panel at OGTT Visit

Next, we examined the methylation patterns of early pregnancy 10-CpG panel using EWAS data measured at the OGTT visit. Only two CpGs (cg15574312 and cg26120476) remained significantly differentially methylated in ADS compared to controls and exhibited a similar directional change to that observed in early pregnancy **(Figure 4a)**.

**Figure 4:**
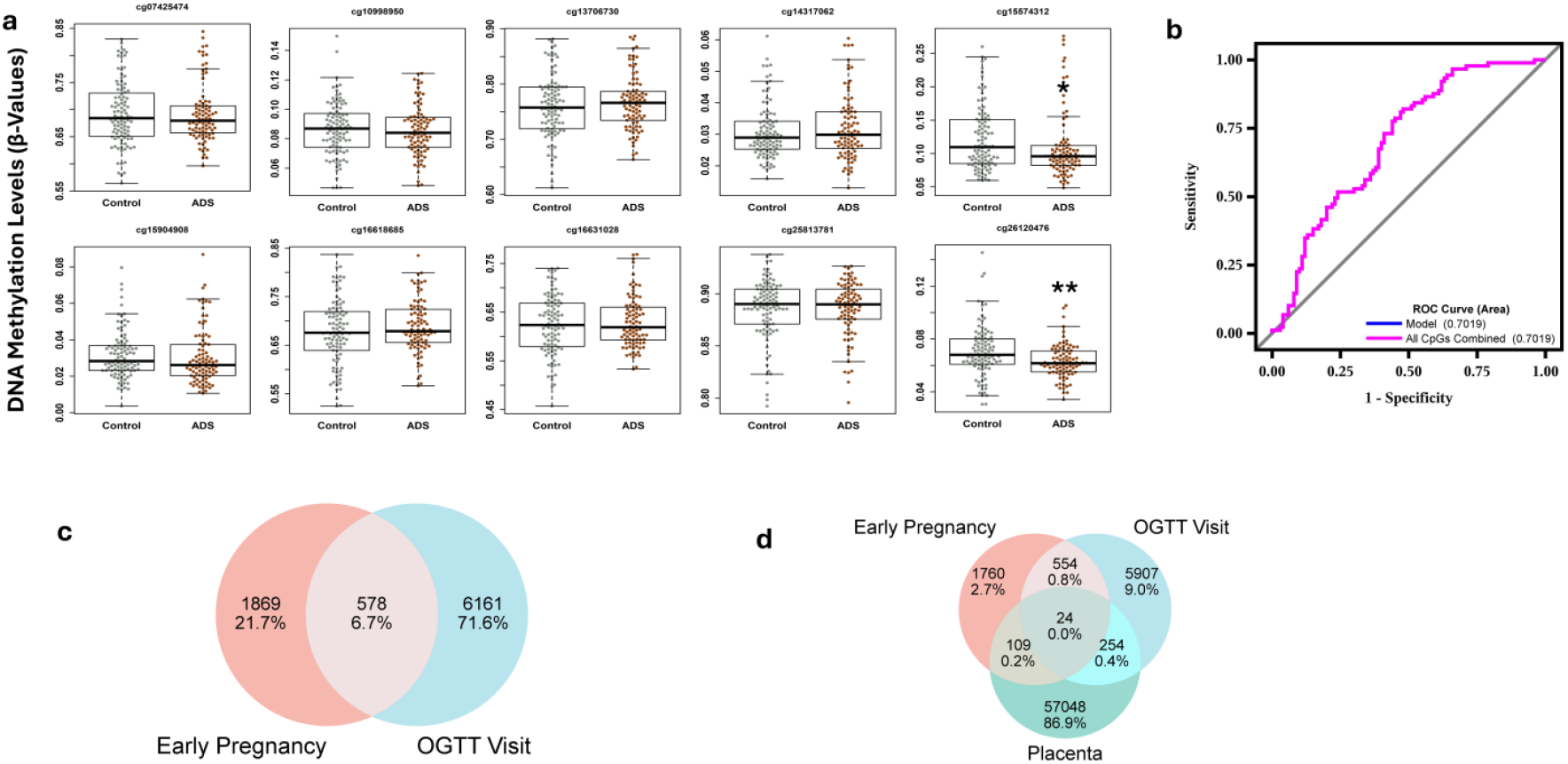
Longitudinal and tissue-level concordance of ADS-associated CpGs. (a) DNA methylation levels of the 10 CpGs at the OGTT visit in relation to ADS and overlap of differentially methylated positions between mADS andblood (early pregnancy and OGTT visit) and placental tissue. Bar graphs represent mean ± SD. *p < 0.05, **p < 0.001 versus controls. Performance of the early-pregnancy 10-CpG panel in discriminating ADS from controls at the time of diagnosis (OGTT visit). (c) Venn diagram showing common DMPs between early pregnancy and OGTT visit blood samples. (d) Venn diagram illustrating shared DMPs across early pregnancy blood, OGTT visit blood, and placental tissue.

To assess the discriminative ability of the early pregnancy 10 CpG panel at the time of diagnosis (during the OGTT visit), ROC analysis was performed, which showed moderate discrimination (AUC = 70%) between ADS and controls **(Figure 4b)**. These results indicate that, two CpGs are consistently found similar methylation pattern throughout different stages of pregnancy and early pregnancy methylation signatures still had a moderate discriminative ability at the time of ADS diagnosis (OGTT visit).

### Similarity of DMPs between maternal blood (early pregnancy and OGTT visit) and placental tissue

We further investigated the similarity of the DMPs between early pregnancy, OGTT visit, and placental tissue. A total of 578 CpGs (6.7%) (FDR<0.05) were differentially methylated between early pregnancy and OGTT visit in maternal blood **(Figure 4c)**. There were 109 common DMPs between early pregnancy and placenta and 254 common DMPs between the OGTT visit and the placenta. Importantly, only 24 DMPs were commonly found in all three datasets (early pregnancy, OGTT, and placenta) **(Figure 4d)**.

Further, we analysed the methylation levels of the 10 CpG panel identified during early pregnancy using the placental EWAS data. There were no significant differences between ADS and control were observed in the placenta **(Figure 5a)**. These findings suggest that some DMPs were shared between the maternal blood and the placental tissue; however, the early pregnancy 10-CpG panel does not show a consistent methylation change in the placenta, suggesting maternal-specific rather than placental-driven regulation of ADS.

**Figure 5:**
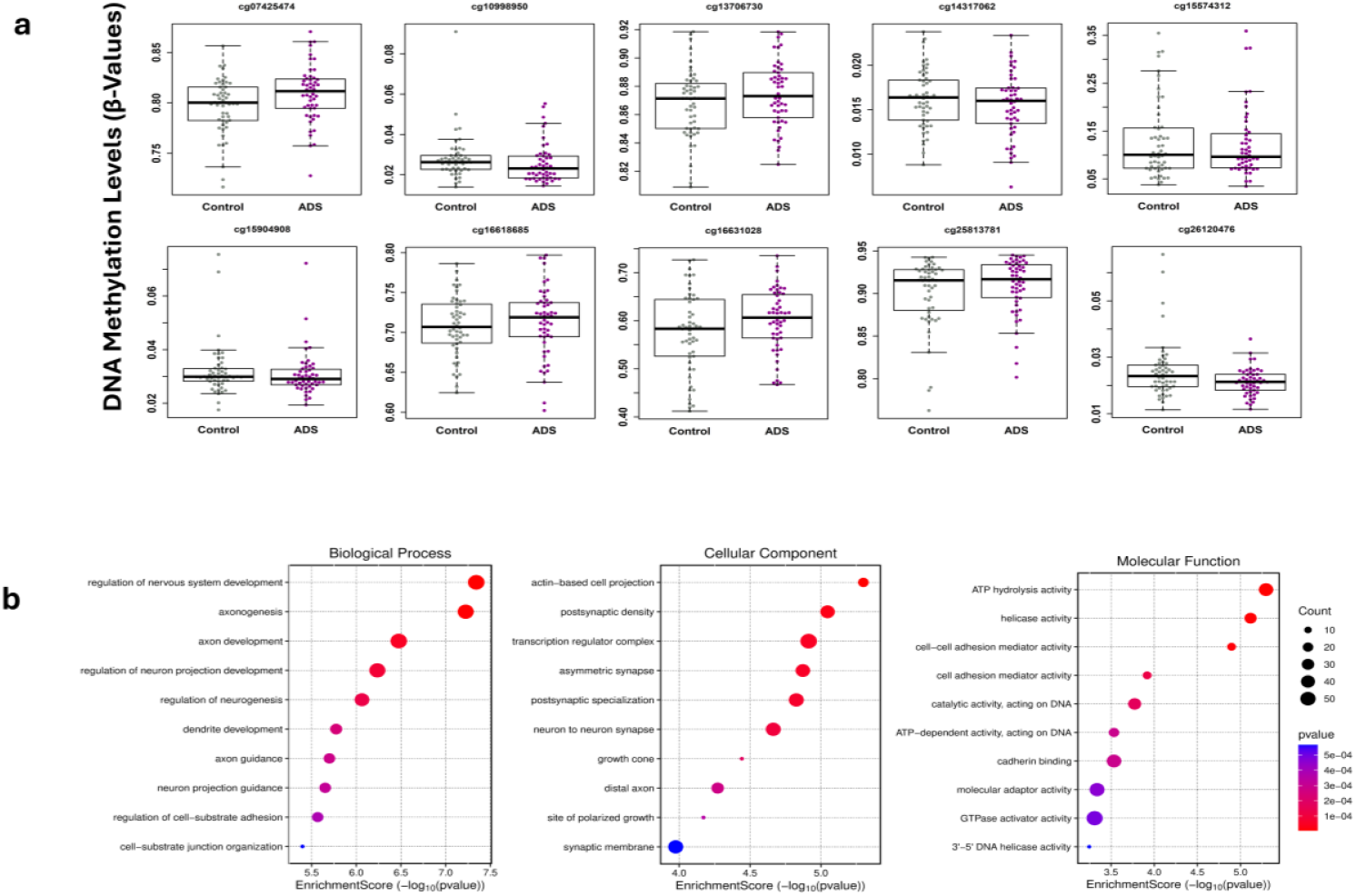
Placental DNA methylation and functional enrichment of ADS-associated CpGs. (a) DNA methylation levels of the 10 top-performing CpGs in placental tissue from controls and ADS cases, presented as mean ± SD. (b) Bar plots showing Gene Ontology (GO) enrichment analysis of genes annotated to DMPs, highlighting the top 10 terms for biological process, molecular function, and cellular component categories.

### Functional Enrichment Analysis of ADS-Associated DMPs

To identify the possible biological mechanisms underlying ADS, we conducted Gene Ontology (GO) and KEGG pathway enrichment analysis for 2,447 DMPs (FDR < 0.05) and their annotated 718 genes. There were three GO categories, Biological Processes (BP), Cellular Components (CC), and Molecular Functions (MF). The top 10 GO enrichments in BP categories, included regulation of nervous system development, axonogenesis, regulation of neurogenesis, dendrite development, axon guidance and neuron projection guidance. In CC category were actin-based cell projection, postsynaptic density, transcription regulator complex, asymmetric synapse, postsynaptic specialization and neuron-to-neuron synapse. In the case of MF, ATP hydrolysis activity, helicase activity, cell-cell adhesion mediator activity, catalytic activity against DNA and ATP-dependent DNA-related activity **(Figure 5b)**.

KEGG pathway analysis of 718 genes revealed that there was a significant enrichment of various pathways such as the sphingolipid signalling, Hippo signalling, Notch signalling, Wnt signalling, insulin resistance, Th1/Th2 cell differentiation, axon guidance, neuroactive ligand-receptor interaction, neurotrophin signalling, cAMP signalling, Alzheimer disease, AMPK signalling, longevity-regulating pathway, calcium signalling and insulin signalling **(Table 4)**. These findings suggests that DNA methylation alterations associated with ADS are over enriched in neurodevelopment, synaptic activity, and metabolic signalling pathways, indicating possible molecular pathways of ADS.

**Table 4:** KEGG pathway enrichment analysis of differentially methylated CpGs and their annotated genes.

| **ID** | **Description** | **p value** | **p adjusted** | **q value** | **Enrichment Score** | **Fold Enrichment** |
| --- | --- | --- | --- | --- | --- | --- |
| hsa04071 | Sphingolipid signalling pathway | 1.3708E-05 | 0.003 | 0.002 | 4.86 | 2.57 |
| hsa04390 | Hippo signalling pathway | 1.8803E-05 | 0.003 | 0.002 | 4.72 | 2.33 |
| hsa04330 | Notch signalling pathway | 4.77433E-05 | 0.003 | 0.002 | 4.32 | 3.16 |
| hsa04310 | Wnt signalling pathway | 4.9679E-05 | 0.003 | 0.002 | 4.30 | 2.18 |
| hsa04931 | Insulin resistance | 0.000542059 | 0.015 | 0.011 | 3.26 | 2.28 |
| hsa04658 | Th1 and Th2 cell differentiation | 0.000596634 | 0.015 | 0.011 | 3.22 | 2.39 |
| hsa04360 | Axon guidance | 0.000744232 | 0.017 | 0.013 | 3.12 | 1.92 |
| hsa04082 | Neuroactive ligand signalling | 0.00116645 | 0.025 | 0.020 | 2.93 | 1.84 |
| hsa05017 | Spinocerebellar ataxia | 0.001311231 | 0.027 | 0.021 | 2.88 | 1.99 |
| hsa04722 | Neurotrophins signalling pathway | 0.001789584 | 0.033 | 0.025 | 2.74 | 2.07 |
| hsa04024 | cAMP signalling pathway | 0.002019162 | 0.034 | 0.026 | 2.69 | 1.73 |
| hsa05010 | Alzheimer disease | 0.002057261 | 0.034 | 0.026 | 2.68 | 1.53 |
| hsa04152 | AMPK signalling pathway | 0.00217723 | 0.034 | 0.026 | 2.66 | 2.03 |
| hsa04211 | Longevity regulating pathway | 0.003171911 | 0.040 | 0.031 | 2.49 | 2.18 |
| hsa04020 | Calcium signalling pathway | 0.003367887 | 0.040 | 0.031 | 2.47 | 1.64 |
| hsa04910 | Insulin signalling pathway | 0.003992859 | 0.044 | 0.034 | 2.39 | 1.89 |
| hsa04144 | Endocytosis | 0.005512637 | 0.052 | 0.040 | 2.25 | 1.60 |

## Discussion

The present study yielded significant findings, using a longitudinal design and epigenome-wide DNA methylation profiles, during early pregnancy. We identified 2,447 differentially methylated positions (DMPs) in early pregnancy blood samples, associated with the subsequent development of ADS, indicating that epigenetic changes occur before the onset of the clinical manifestation of depressive symptoms. Second, using machine-learning feature selection, we identified a robust biomarker panel of 10-CpGs that strongly predicted ADS with high accuracy and this prediction was significant even after adjusting for maternal risk factors. Third, the MRS constructed based on this CpG panel significantly outperformed the traditional maternal risk factors, highlighting the additional and independent contribution of the epigenetic signatures for early risk stratification. Lastly, beyond prediction, functional enrichment analyses identified ADS-related methylation alterations were significantly enriched in neurodevelopmental, synaptic, immune, and metabolic pathways, supporting biological plausibility. To our knowledge, this is one of the first studies from a low- and middle-income country reporting the feasibility that epigenetic markers could be used to predict ADS during early pregnancy.

Earlier epigenetic studies have shown that the epigenetic alterations in genes involved in glucocorticoid signalling (e.g., NR3C1), neurotrophin pathways (e.g., BDNF), and immune regulation, were associated with antenatal and postpartum depression (Kundakovic and Jaric, 2017; Palma-Gudiel et al., 2015). Viuff et al. (2018) analysed cord blood DNA methylation in 844 mother-child pairs in Avon Longitudinal Study of Parents and Children (ALSPAC) cohort and found two CpG sites linked to mid-pregnancy depression and 39 differentially methylated regions (DMRs) during gestation (Viuff et al., 2018). Similarly, King et al. (2017) found that mothers with persistent perinatal depression had significantly higher methylation in the oxytocin receptor (OXTR) gene and lower methylation in the adjacent intergenic area of the vasopressin (AVP) gene (King et al., 2017). U.S. cohort evidence further suggests that DNA methylation at a specific locus could be a predictor of PPD. Guintivano et al. (2014) showed that the methylation changes in TTC9B and HP1BP3 were notable predictors of PPD, which were subsequently validated in two independent cohorts by Payne et al. (2020) (Guintivano et al., 2014; Payne et al., 2020). These studies, however, could not determine the time precedence or predictive utility.

We did not recap these findings, which could be attributed to population-scale methylation heterogeneity, since DNA methylation patterns have been shown to be ancestry and ethnicity-specific (Elliott et al., 2022; Song et al., 2021). Conversely, the present study uniquely reported that the early pregnancy methylation panel measured before the clinical manifestations is a strong predictor of subsequent ADS. Moreover, the use of machine-learning approaches allowed us to shift our attention beyond single-CpG associations to clinically useful biomarker panels. The predictive performance observed here is higher than that in many prior EWAS studies of depression, underscoring the importance of using epigenome-wide information, along with advanced analytical models. We also observed that the maternal risk factors alone did not predict well, but the addition of epigenetic data significantly improved prediction, indicating the potential of precision medicine methods in perinatal mental health.

A central question in epigenetic studies of depression is whether observed DNA methylation differences merely correlate with depressive symptoms or whether they may participate in biological pathways that contribute to disease vulnerability. While causality cannot be definitively established, the functional relevance of genes annotated to the 10-CpG biomarker panel supports the hypothesis that these methylation changes may participate in molecular pathways that increase susceptibility to ADS. Importantly, cg07425474 and cg14317062 emerged as the most influential CpGs in the predictive model. The gene annotated to cg07425474 has been implicated to synaptic maturation and neuronal circuit formation, and the impact of its expression has been experimentally shown to cause abnormal synaptic pruning and behavioural symptoms of mood disorders in animal models (He et al., 2025). We observed a higher methylation pattern at this locus in women who subsequently developed ADS, may plausibly contribute to reduced gene expression, thereby impairing synaptic plasticity and emotional regulation during pregnancy. These processes align closely with neuroplasticity-based depression theories, which argue that altered neuronal plasticity and changes in synaptic connectivity underlie vulnerability to mood disorders (Player et al., 2013; Wainwright and Galea, 2013).

Remaining CpGs in the panel were mapped to genes involved in synaptic organisation, axon guidance and dendritic development. These genes could be epigenetically regulated to modulate neuronal signalling efficiency and stress responsivity, processes that are important in maintaining emotional stability during pregnancy (Duan et al., 2017; Wang et al., 2023; Wenzel et al., 2025). Emerging studies suggest that peripheral epigenetic signatures may be indicative of systemic neurobiological conditions, especially in stress- and immune-related pathways (Houtepen et al., 2016; Uddin et al., 2010). Thus, these CpGs may act as accessible peripheral indicators of central nervous system vulnerability rather than passive correlates.

Notably, the CpG panel exhibited both negative and positive correlations with ADS risk, suggesting possible protective or resilience-associated epigenetic patterns. Hypomethylation or hypermethylation at these loci could alter the gene expression of neuronal resilience, metabolic homeostasis or anti-inflammatory signalling genes. This bidirectional pattern of hyper- and hypomethylation supports a causal framework in which antenatal depression risk arises from an imbalance between epigenetically mediated vulnerability and protective pathways, rather than uniform dysregulation across the genome. This systems-level interpretation is further supported by the strong performance of the MRS, which integrates small methylation differences across multiple biologically interconnected genes. The MRS markedly outperformed the maternal risk factors and improved prediction when combined with clinical variables, consistent with a cumulative biological risk model. Collectively, these gene-level findings suggest that the discovered methylation signatures do not merely serve as statistical predictors but may represent early molecular signatures of perturbed neuroimmune and synaptic control underlying depression. Our findings, therefore, provide proof of concept and can be used for future mechanistic studies on the possible role of these methylation signatures in the pathogenesis of ADS in various ethnic groups. Additionally, the identified CpG panel can be useful in screening individuals at high risk. However, further validation in larger, independent cohorts and different ethnicities is needed to confirm these results.

An important aspect of the present study is the comparison of methylation patterns between maternal blood at different visits and placental tissue among the same individuals. Although a subset of DMPs overlapped between maternal blood and placenta, only a few CpGs were found to be common in the early pregnancy, OGTT visit, and placental samples. Notably, the 10-CpG predictive panel identified in early pregnancy blood did not exhibit consistent methylation patterns in placental tissue. This indicates that the observed methylation pattern primarily reflects maternal systemic or neuroimmune processes rather than the placental pathology. This is in line with the prior studies demonstrating limited cross-tissue concordance of methylation alterations related to depression, highlighting the tissue-specificity of epigenetic regulation (Lowe et al., 2015). Our results strengthen the clinical feasibility of maternal blood-based biomarkers and support the interpretation that maternal biological vulnerability is a significant determinant of ADS risk.

Functional enrichment analyses of ADS-associated DMPs strengthened these mechanistic interpretations, which showed overrepresentation of pathways related to synaptic organization, neurotrophin signaling, immune regulation, nervous system development and metabolic signaling. KEGG pathway analysis of DMPs and their annotated genes showed that there was significant enrichment of Wnt, Notch, AMPK and insulin signalling pathways molecular cascades widely involved in mood regulation and metabolic functioning. In line with this, previous studies reported that bipolar disorder (BD) and schizophrenia (SCZ) have altered Wnt signalling with a reduction in canonical (β-catenin–dependent) activity and evidence of increased non-canonical activity (Hoseth et al., 2018a). The role of notch signalling has been linked to psychiatric disorders and increased plasma concentrations of Notch ligands and decreased expression of genes related to the pathway (e.g. higher RFNG and KAT2B, lower PSEN1 and CREBBP) in SCZ and BD compared to healthy controls have also been reported (Hoseth et al., 2018b; Zhang et al., 2025). The enrichment of insulin and AMPK signaling pathways further suggests shared biological mechanisms linking metabolic dysregulation and depressive vulnerability during pregnancy. These results suggest the biological plausibility of our pathway findings, indicating that dysregulation in interconnected neurodevelopmental and metabolic signalling pathways may contribute to susceptibility to antenatal depression.

This study has notable strengths. First, the longitudinal design enables the identification of epigenetic changes that precede the onset of depressive symptoms, thereby strengthening causal inference. Second, the exclusion of women with pre-existing psychiatric illness minimises confounding by pre-existing depression or psychotropic medication use. Third, the combination of EWAS with powerful machine-learning techniques, such as bootstrap validation and SHAP-based interpretability, enhances the reliability and translational relevance of the identified biomarkers. Fourth, the study population from India addresses a major gap in the literature, as most epigenetic studies of antenatal depression have been conducted in Western populations. While causality cannot be definitively established, this temporal ordering, combined with biologically plausible gene annotations, is consistent with a contributory rather than purely consequential role of the identified epigenetic alterations.

However, limitations need to be acknowledged. The sample size, while adequate for machine-learning–based prediction, remains modest for EWAS studies, and replication in larger, independent cohorts is warranted. The use of peripheral blood limits direct inference about brain-specific epigenetic changes, although prior evidence suggests partial concordance between blood and central nervous system methylation for stress-related pathways (Edgar et al., 2017). Additionally, depressive symptoms were assessed using the PHQ-9 rather than diagnostic interviews, which may have led to some degree of misclassification. Finally, although we adjusted for major maternal confounders, residual confounding by unmeasured psychosocial stressors cannot be entirely excluded.

In conclusion, this study identifies early pregnancy DNA methylation signatures that predict antenatal depressive symptoms before clinical onset, addressing a critical gap in perinatal mental health screening. The 10-CpG biomarker panel captures biologically relevant neuroimmune and metabolic pathways, substantially improving risk prediction beyond clinical factors. These findings support the integration of objective, blood-based epigenetic screening into routine antenatal care, enabling the early identification of mental health issues and the implementation of preventive interventions, particularly within resource-constrained public health systems.

## Acknowledgments/Funding

Dr. Kuppan Gokulakrishnan is a current recipient of a DBT-Wellcome Trust India Alliance Intermediate Clinical & Public health Fellowship (Grant Number IA/CPHI/18/1/503964) and acknowledges the funding received from the DBT-Wellcome Trust India Alliance for this study. STRiDE study was funded by MRC-DBT Newton fund (MRC Newton Fund MR/N006232/1).

## Author’s contributions

KG is the principal investigator of this study and has conceptualized, written, and taken the lead in completing manuscripts through all stages of preparation and submission. MB, PS, RMA, UR and VM have reviewed the project from the conceptualization stage and have contributed to each version of the manuscript. CT, MD, BNS, and UR are members of the research team who participated in the conduct of the study and have contributed to writing the manuscript. CT performed experiments. KG, KS, BB, VU and CT were involved in data analysis. All authors have contributed to the article critically for intellectual content and have provided final approval of the version to be published.

## Data availability statement

The data that support the findings of this study are available from the corresponding author, [KG], upon reasonable request.

