## Supplementary figures and images for "Early Pregnancy DNA Methylation Signatures as Predictors of Antenatal Depressive Symptoms: A longitudinal study of DNA methylation changes"

### Supplemental Figure

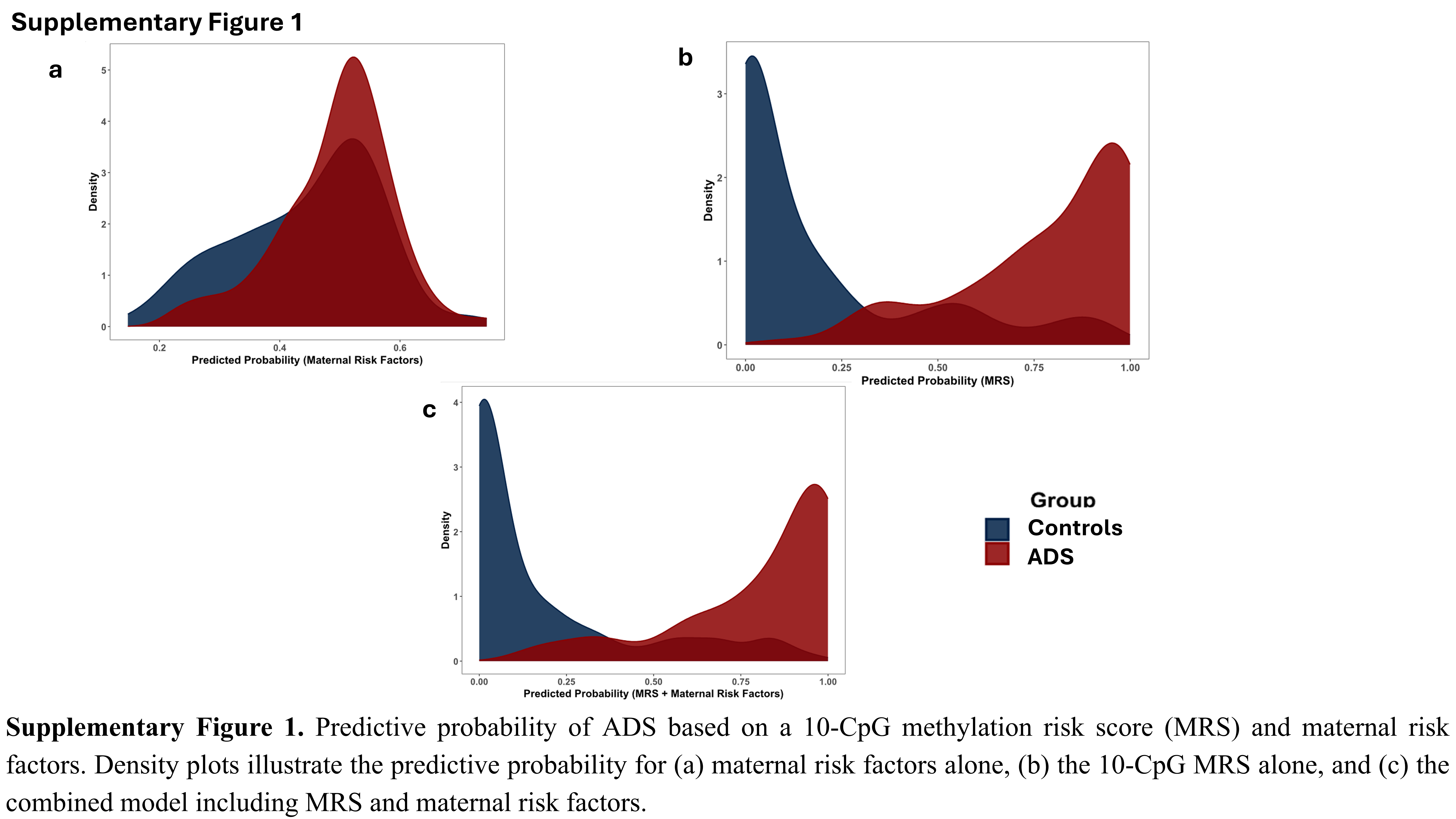
