## Supplemental table 1 for "Early Pregnancy DNA Methylation Signatures as Predictors of Antenatal Depressive Symptoms: A longitudinal study of DNA methylation changes"

**Supplementary Table 1: Machine learning data analysis framework**

A dataset consisting of CpGs from various feature extraction techniques was used as the input dataset.

Panel size was fixed as max 10.

The number of iterations was fixed as 10 iterations.

For each iteration:

- Randomly split the data into 67% training set and 33% testing set.
- Stepwise Predictor Selection with Parameter Tuning (includes 10 rounds)
  - For each round
    - Take one CpG at a time in the model to tune the parameters, using 10-fold cross-validation repeating over 5 times. Select the first CpG in the panel with the highest AUC.
    - For each remaining predictor:
      - Add a predictor at a time in combination with the current set of selected predictors.
      - Train a model using the current set of predictors on the training set with parameter tuning.

Set up cross-validation and tuning grid.

Train the model with cross-validation and tune parameters.

- Evaluate the model on the testing set, calculating performance metrics such as AUC, Accuracy, Sensitivity, and Specificity.
- Select Best Predictor
  - Identify the predictor that results in the best model performance (highest AUC).
- Update Selected Predictors
  - Add the best predictor to the list of selected predictors.
  - Update the best model and performance metrics if the current model is the best.
- Store Round Results
  - Store the performance metrics and selected predictors for the current round.
