## Supplemental table 2 for "Early Pregnancy DNA Methylation Signatures as Predictors of Antenatal Depressive Symptoms: A longitudinal study of DNA methylation changes"

**Supplementary Table 2:** ROC analysis of the 10 CpGs biomarker panel in the whole data set.

|  | **Panel of 10 CpGs** |
| --- | --- |
| **AUC** | 0.94 |
| **Sensitivity (%)** | 87 |
| **Specificity (%)** | 84 |
| **Accuracy (%)** | 86 |
| **LR Positive** | 5.47 |
| **LR Negative** | 0.15 |
| **PPV** | 0.83 |
| **NPV** | 0.88 |
